# Racial and Ethnic Disparities in Population Level Covid-19 Mortality

**DOI:** 10.1101/2020.05.07.20094250

**Authors:** Cary P. Gross, Utibe R. Essien, Saamir Pasha, Jacob R Gross, Shi-yi Wang, Marcella Nunez-Smith

**Affiliations:** National Clinician Scholars Program, Yale School of Medicine, New Haven, CT; Division of General Internal Medicine, University of Pittsburgh School of Medicine; Cancer Outcomes, Public Policy and Effectiveness Research (COPPER) Center, Yale Cancer Center, New Haven, CT, USA; School of Arts & Sciences, Tufts University, Medford, MA; Department of Chronic Disease Epidemiology, Yale School of Public Health, New Haven, CT, USA; Equity Research and Innovation Center, Yale University, New Haven, CT, USA

## Abstract

**Background:** Current reporting of Covid-19 mortality data by race and ethnicity across the United States could bias our understanding of population-mortality disparities. Moreover, stark differences in age distribution by race and ethnicity groups are seldom accounted for in analyses.

**Methods:** To address these gaps, we conducted a cross-sectional study using publicly-reported Covid-19 mortality data to assess the quality of race and ethnicity data (Black, Latinx, white), and estimated age-adjusted disparities using a random effects meta-analytic approach.

**Results:** We found only 28 states, and NYC, reported race and ethnicity-stratified Covid-19 mortality along with large variation in the percent of missing race and ethnicity data by state. Aggregated relative risk of death estimates for Black compared to the white population was 3.57 (95% CI: 2.84-4.48). Similarly, Latinx population displayed 1.88 (95% CI: 1.61-2.19) times higher risk of death than white patients.

**Discussion:** In states providing race and ethnicity data, we identified significant population-level Covid-19 mortality disparities. We demonstrated the importance of adjusting for age differences across population groups to prevent underestimating disparities in younger population groups. The availability of high-quality and comprehensive race and ethnicity data is necessary to address factors contributing to inequity in Covid-19 mortality.

## Background

Alarming racial and ethnic disparities have been reported regarding risk of Covid-19 infection, access to testing, and adverse outcomes.^1^ Such disparities may be attributed to the disproportionate burden experienced by people of color regarding comorbidity, at-risk employment, unstable housing, incarceration, and decreased access to healthcare and medical resources. Unfortunately, as of mid-April, only half of states were reporting Covid-19 mortality by race and ethnicity, while Centers for Disease Control and Prevention (CDC) Covid-19 data are missing race and ethnicity on 65% of patients.^2,3^

These incomplete data impact national estimates: the CDC data^4^, suggests that white patients represent a higher proportion of Covid-19 diagnoses than their representation in the general population.^5^ Yet data derived from specific regions that report race and ethnicity of Covid-19 decedents, show that Black patients are dying at a much higher rate than their population share. Other work has shown that counties with a disproportionately high Black population accounted for over half of all Covid-19 deaths while making up just 35% of the population.^6^

Among states that are reporting race and ethnicity data, the quality of the data and the magnitude of population-mortality disparities remain unclear. Further, the profound differences in population age distribution across racial and ethnic groups are infrequently incorporated into analyses. Given the strong relation between age and risk of Covid-related mortality, and the younger age distributions of the Black and Latinx populations, these unadjusted data can result in substantial underestimates of disparities. To address these gaps, we therefore evaluated the quality of race and ethnicity reporting in Covid-19 mortality data across states, and estimated age-adjusted disparities in population mortality rates.

## Methods

We conducted a cross-sectional study using publicly-reported Covid-19 mortality data available on state websites^2,7^ as of April 21, 2020. We focused on Black, Latinx and white populations as they are the largest racial and ethnic groups represented in these data. State and county population demographics were obtained from 2017 United States Census estimates data^8^. New York State and New York City (NYC) report their data separately and were considered as distinct units for this study.

We used indirect standardization to assess the population mortality rate across racial and ethnic groups, accounting for the younger age distributions of the Latinx and Black populations.^8,9^ First, we determined the relative risk of Covid-19 death at the national level across age groups (<45, 45-54, 55-64, 65-74, and 75+ years old), based on CDC data.^5^ We then used these age-related relative risk data to estimate the relevant mortality rates for patients in each age/race/ethnic group by state, based on each state’s observed race and ethnicity-specific Covid-19 mortality rate. We used these rates to estimate the number of deaths that would be observed if each group had the same age distribution as the white population in that state, estimating the relative risk and 95% confidence interval (95% CI) for each state and using a random effects meta-analytic approach to derive a reliable summary estimate of disparities.^10^ Finally, we conducted a meta-regression analysis to assess the association between state-level racial disparities (i.e. Black vs white mortality risk ratio, RR) and three state characteristics: percent of decedents missing race/ethnicity data, percent urban, and percent of state population Black/Latinx.

## Results

We found that 28 states, and NYC, reported race and ethnicity-stratified Covid-19 mortality. There was substantial variation in the percent with missing race and ethnicity data, with only eight states missing such data on <5% of decedents (Figure 1). There was substantial variation in the association between Black race and mortality across states (Figure 2a). In 22 states (plus NYC), the risk of Covid-19 associated death was significantly higher for the Black than the white population, ranging as high as 18-fold higher in Wisconsin. In one state (Pennsylvania), the Black population had a significantly lower population mortality (RR: 0.44; 95% CI: 0.24-0.69). When aggregating the data from all states (and NYC), the relative risk of death for Black compared to the white population was 3.57 (95% CI: 2.84-4.48).

**Figure 1.**
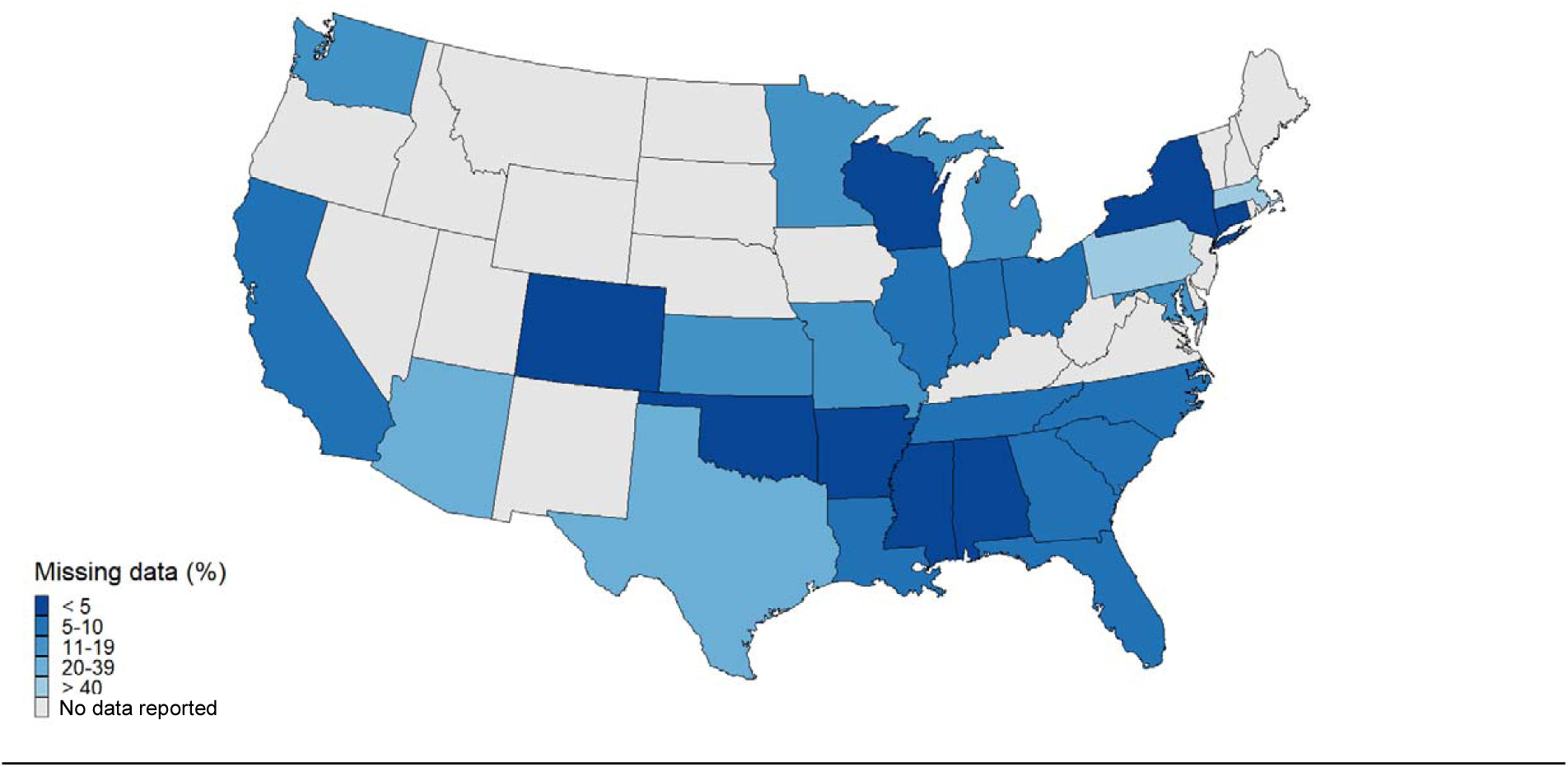
Percent race/ethnicity information missing in COVID death data by State, April 21,2020

**Figure 2a:**
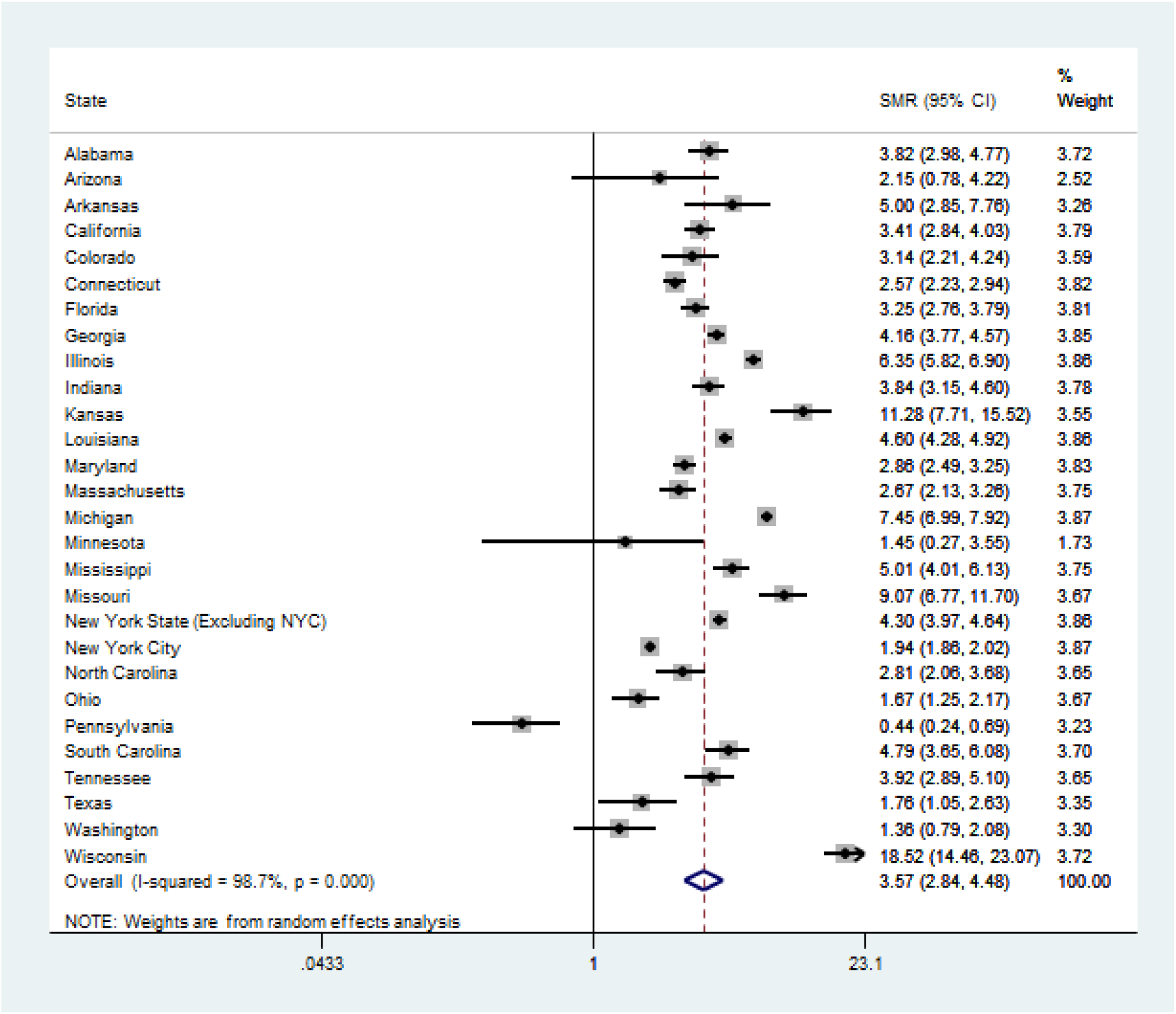
Covid-19 Related Mortality in Black vs White Populations by State^*^

Findings were similar for the Latinx population, which experienced an 88% higher risk of death than white patients (RR for Latinx vs White: 1.88: 1.61-2.19). In 12 states, as well as NYC, Latinx people had a significantly higher risk of Covid-related mortality than white people (Figure 2b). There were no states in which the Latinx population had a significantly lower mortality.

**Figure 2b.**
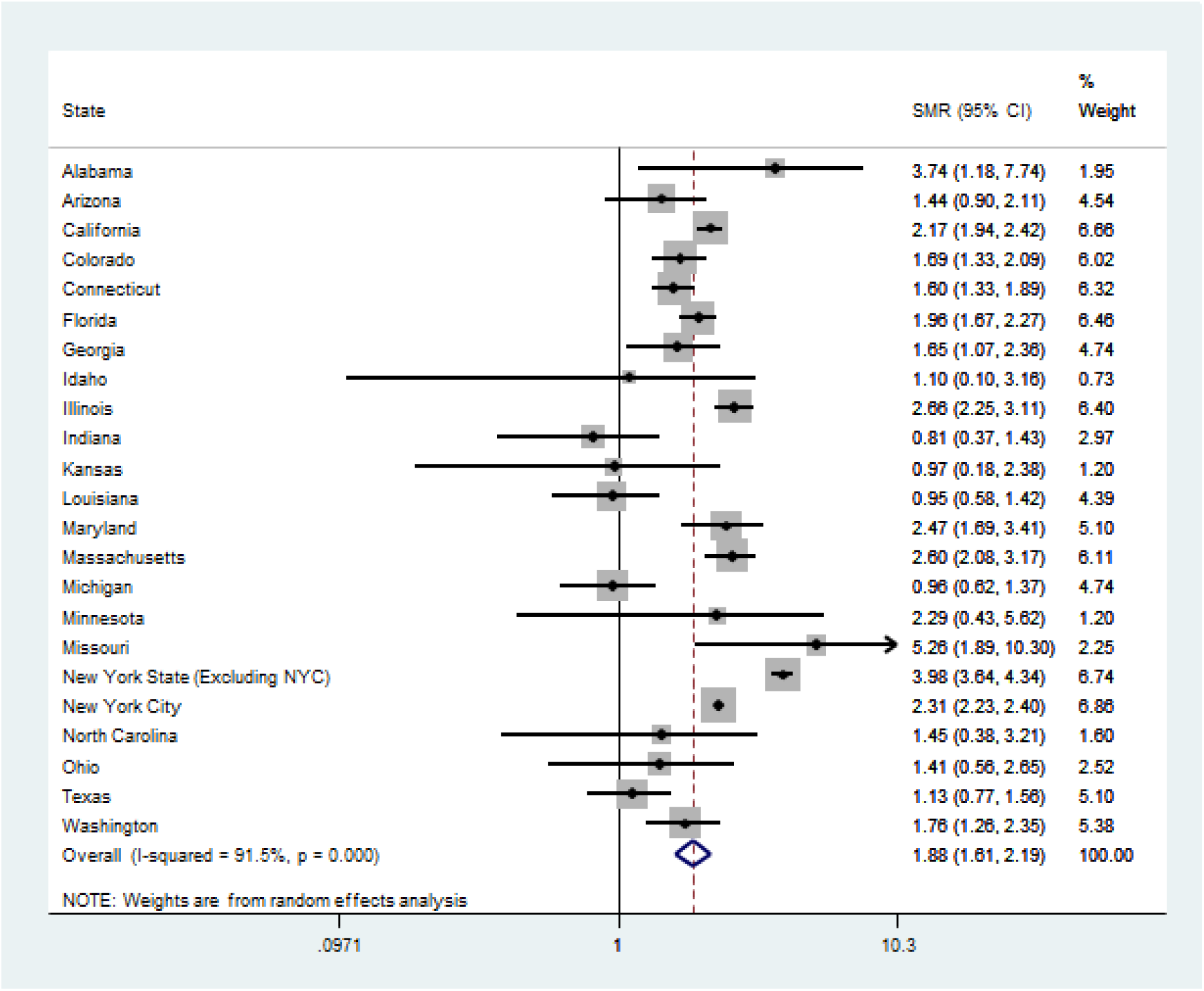
Covid-19 Related Population Mortality in Latinx vs White Populations by State^*^

In both analyses, the I-square was >90%, suggesting substantial heterogeneity across states in the magnitude of disparities. For the meta-regression analysis, we found that the percent of the population that was Black or Latinx, or percent urban, was not associated with the magnitude of disparities. However, states with higher proportion of deaths that were missing race data tended to have lower disparities (i.e, a lower Black vs white effect size; P=0.047)

## Discussion

We found several findings that build upon prior work related to disparities in the Covid-19 pandemic in important ways. First, among states that are reporting race and ethnicity mortality data, we found a strong relation between Black race, Latinx ethnicity, and population-level Covid-19 mortality. Notably, we also found that the magnitude of these Covid-19 disparities varied substantially across states. While some states do not have demonstrable disparities, minorities in other states face 5 or 10-fold or higher risk of death than their white counterparts. Our findings also underscore the importance of adjusting for age differences across population groups, given that unadjusted mortality rates will underestimate disparities for younger population groups. In many states, the crude mortality rate for the Latinx population is lower than the white population; a disparity that reverses after considering age. Additionally, our study is the first to our knowledge to report the substantial variability in missingness of race/ethnicity data across states. This is crucial to correct, because poor quality data intrinsically limits our ability to conduct rigorous research that can provide critical insights on how best to target and evaluate public health and clinical interventions to reduce health disparities.

It is important to note that it is unclear whether underreporting of death related to Covd-19 occurs unequally across racial/ethnic groups. Future work should explore Covid-19 outcomes in other racial groups, as well as variability in the reporting of other relevant Covid-19 data including access to testing and infection rates. Additionally, we applied age-related mortality risks, derived from national data, to individual state data, due to the paucity of race/ethnicity/age-specific data for each state. However, when we compared our approach to a state that does report its own age-adjusted rates by race group, we found remarkably similar results: the relative risk for Black vs white age-adjusted mortality as estimated by New York State was 4.17 (compared to our estimate of 4.30). Finally, it is unclear how race and ethnicity are determined by each state. Ideally, the method should focus on how patients self-identified, which can be challenging during a pandemic, particularly in areas where coroners/mortuary staff are overwhelmed

Public health officials, healthcare systems, and policymakers should work together to improve the availability of high-quality and comprehensive race and ethnicity data, and to thoroughly investigate and mitigate factors that contribute to inequity in Covid-19 mortality.

<u>Figure 2 Legend:</u> *New York City is included as a separate region, as these data are reported distinctly from New York State data; SMR: Standardized Mortality Ratio for Black or Latinx in comparison to white population.

## Data Availability

All data retrieved from publicly available sources

https://www.cdc.gov/nchs/nvss/vsrr/covid_weekly/

https://www.cdc.gov/coronavirus/2019-ncov/cases-updates/cases-in-us.html

https://ehe.amfar.org/disparities

https://covidtracking.com/data

https://www.census.gov/newsroom/press-kits/2018/estimates-characteristics.html

